# The emergence of azithromycin-resistant *Salmonella* Typhi in Nepal

**DOI:** 10.1101/2020.08.07.20166389

**Authors:** Pham Thanh Duy, Sabina Dongol, Abhishek Giri, To Nguyen Thi Nguyen, Thanh Ngoc Dan Ho, Quynh Pham Nhu Nguyen, Trung Duc Pham, Guy E. Thwaites, Buddha Basnyat, Stephen Baker, Maia A. Rabaa, Abhilasha Karkey

**Author notes:** Joint senior authors (Maia A. Rabaa, Abhilasha Karkey). Corresponding author: Dr Pham Thanh Duy, Molecular Epidemiology Group, Oxford University Clinical Research Unit, Vietnam.

## Abstract

Typhoid fever remains a significant cause of morbidity and mortality in Asia and Africa. The emergence of azithromycin resistance in South Asia is concerning, as azithromycin is one of the last effective oral drugs for treating typhoid. In mid-2019, three azithromycin-resistant (Azith^R^) S. Typhi isolates were isolated from typhoid fever patients attending Patan Hospital, Kathmandu, Nepal. These organisms were whole genome sequenced and compared with a global collection. We found that the three Azith^R^ isolates belonged to the H58 lineage and were genetically identical; they were distantly related to contemporaneous *S*. Typhi from Nepal and Azith^R^ *S*. Typhi recently described in Bangladesh. Azithromycin resistance was mediated by nonsynonymous mutation in the *acrB* gene (R717L). Clinical information from one patient suggested non-response to azithromycin treatment. Further investigations are needed to evaluate treatment responses to azithromycin, predict Azith^R^ *S*. Typhi’s evolutionary trajectories, and track the transmission of these organisms.

## Main Text

Typhoid fever is a life-threatening systemic infection predominantly caused by *Salmonella enterica* serovar Typhi *(S*. Typhi). Though the disease has been controlled in developed countries, it continues to cause significant morbidity and mortality in resource-poor settings in Asia and Africa. Effective antimicrobial therapy is essential to avoid deaths and serious complications. However, *S*. Typhi has continually evolved resistance to antimicrobials used for its treatment, posing a constant clinical challenge and likely exacerbating disease burden.^1^ Multi-drug resistance (MDR; resistance to chloramphenicol, ampicillin, trimethoprim-sulfamethoxazole) first evolved in *S*. Typhi in the late 1980s, followed by fluoroquinolone resistance in the 1990s.^2^ Third-generation cephalosporins have since been used for typhoid treatment, but the emergence of extensively-drug resistant (XDR; MDR plus resistance to fluoroquinolones and third-generation cephalosporins) *S*. Typhi in Pakistan^3^ has reduced the clinical efficacy of these drugs and raises concerns regarding the imminent spread of untreatable S. Typhi.

Azithromycin is effectively the last remaining oral antimicrobial to treat typhoid fever.^4^ Although azithromycin resistance in *S*. Typhi has rarely been reported, an increasing reliance on this drug over the last decade has led to the emergence of azithromycin-resistant (Azith^R^) S. Typhi in South Asia. A recent study in Bangladesh indicated that azithromycin resistance (MIC >32 μg/ml) in *S*. Typhi was associated with a non-synonymous mutation (R717Q) in the *acrB* gene, which encodes an efflux pump.^5^ There are limited data on clinical responses to azithromycin in Azith^R^ *S*. Typhi-infected patients. Here, we report the genomics, antimicrobial resistance profiles, and phylogenetic relatedness of three Azith^R^ S. Typhi isolates isolated from typhoid fever outpatients visiting a hospital in Nepal. We report clinical manifestations and azithromycin response data for one of the patients.

Patan Hospital (Kathmandu, Nepal) serves ~320,000 outpatients and ~20,000 inpatients annually. Typhoid fever is frequently managed in the outpatient department (OPD) of the hospital and blood culture is routinely performed following routine microbiological procedures when enteric fever is suspected.^6^ Antimicrobial susceptibility testing is performed by a modified Bauer-Kirby disc diffusion, with Etests^®^ to determine MICs (bioMérieux, France); results are interpreted using CLSI guidelines.^7^ In August and September 2019, routine microbiological diagnostic procedures identified three patients with Azith^R^ *S*. Typhi attending the OPD. Clinical information was only available for one of three patients infected with Azith^R^ *S*. Typhi.

Total genomic DNA from *S*. Typhi isolates (including one contemporaneous azithromycin-susceptible (Azith^S^) isolate) was extracted and whole genome sequenced on an Illumina MiSeq platform (raw data deposited in ENA, project PRJEB37899). Sequence data from this study were combined with 1,508 previously published S. Typhi H58 genomes^3,5,8–12^. Single nucleotide polymorphisms (SNPs) were called using previously described methods^11^, resulting in a final set of 3,326 SNPs. A maximum likelihood phylogeny was inferred from the final 3,326 SNP alignment using RAxML (v8.2.8).^13^ Antimicrobial resistance (AMR) genes and plasmid contents of our isolates were determined using SRST2,^14^ with ARG-Annot database^15^ and Plasmidfinder^16^ used as respective reference databases.

On 25 August 2019, a 28-year-old male from Nakkhu (Lalitpur District) presented to the OPD following four days of anorexia and persistent fever despite three days of paracetamol use. General and systemic examinations were normal except for a fever of 38.9°C on presentation. Investigations revealed a haemoglobin count of 13.7 g/dl, total white blood cell count of 8.6 thousand/μl with increased neutrophils (DLC: N-83, L-17), platelet count of 195 thousand/μl, and C-reactive protein of 18.7 mg/dl. Urine microscopy and analysis were normal and blood culture was performed. A clinical diagnosis of enteric fever was made and oral azithromycin (1g once/day) was administered. The patient was asked to return for the blood culture reports after 72 hours. On day 2 of culture, his blood culture was positive for *S*. Typhi, which was found to be Azith^R^ (6mm zone of inhibition on disc diffusion, MIC >256 μg/ml). The patient did not return for the scheduled 72-hour follow-up, but was traced on day seven of treatment. On day seven, the patient reported a fever of 38.3°C lasting for two days. Physical examination showed no abnormalities, but laboratory examinations were repeated due to the previous blood culture results. Repeat laboratory analysis showed a haemoglobin count of 13.5 g/dl, total white blood cell count of 6.9 thousand/μl, continued increased neutrophils (DLC: N-77, L-23), platelet count of 219 thousand/μl, and C-reactive protein of 23 mg/dl. A repeat blood culture was performed and an Azith^R^ *S*. Typhi was again isolated. The patient was admitted and administered intravenous ceftriaxone. The patient became afebrile after 48 hours of ceftriaxone treatment and was discharged after 7-days of intravenous ceftriaxone. Two additional Azith^R^ *S*. Typhi isolates displaying identical resistance phenotypes were identified from a 53-year-old female from Nakkhu, Lalitpur (25 ^th^ August 2019) and a 26-year-old male from Ramshatol, Ramechhap (9^th^ September 2019); no clinical information was available for these patients.

Genomic and phylogenetic analyses showed that these Azith^R^ S. Typhi isolates and the contemporaneous Azith^S^ isolate from this study belonged to the H58 lineage. The three Azith^R^ isolates were genetically identical and differed from the Azith^S^ isolate by 42 SNPs; the three Nepali Azith^R^ isolates were also distantly related to all other H58 isolates from the global collection (Figure 1), and were not linked to the Azith^R^ H58 isolates recently reported in Bangladesh.^5^ Given that most previously published Nepali H58 isolates grouped within sub-lineage II (including the Azith^S^ isolate from this study), the Nepali Azith^R^ isolates likely evolved from an older H58 variant, with a phylogenetic distance between the Azith^R^ isolates and H58 sub-lineage II of at least 27 SNPs.

Regarding AMR, none of the Azith^R^ H58 isolates carried an acquired AMR gene, but harboured a nonsynonymous mutation in the *acrB* gene (STY0519), changing arginine (R) to leucine (L) at codon 717. R717L and R717Q mutations in *acrB* have been found to confer Azith^R^ in Bangladeshi S. Paratyphi A and S. Typhi, respectively.^5^ Furthermore, the Nepali Azith^R^ H58 isolates exhibited double mutations in the *gyrA* gene (S83F, D87G), leading to reduced fluoroquinolone susceptibility; no *parC* mutations were observed.

**Figure 1:**
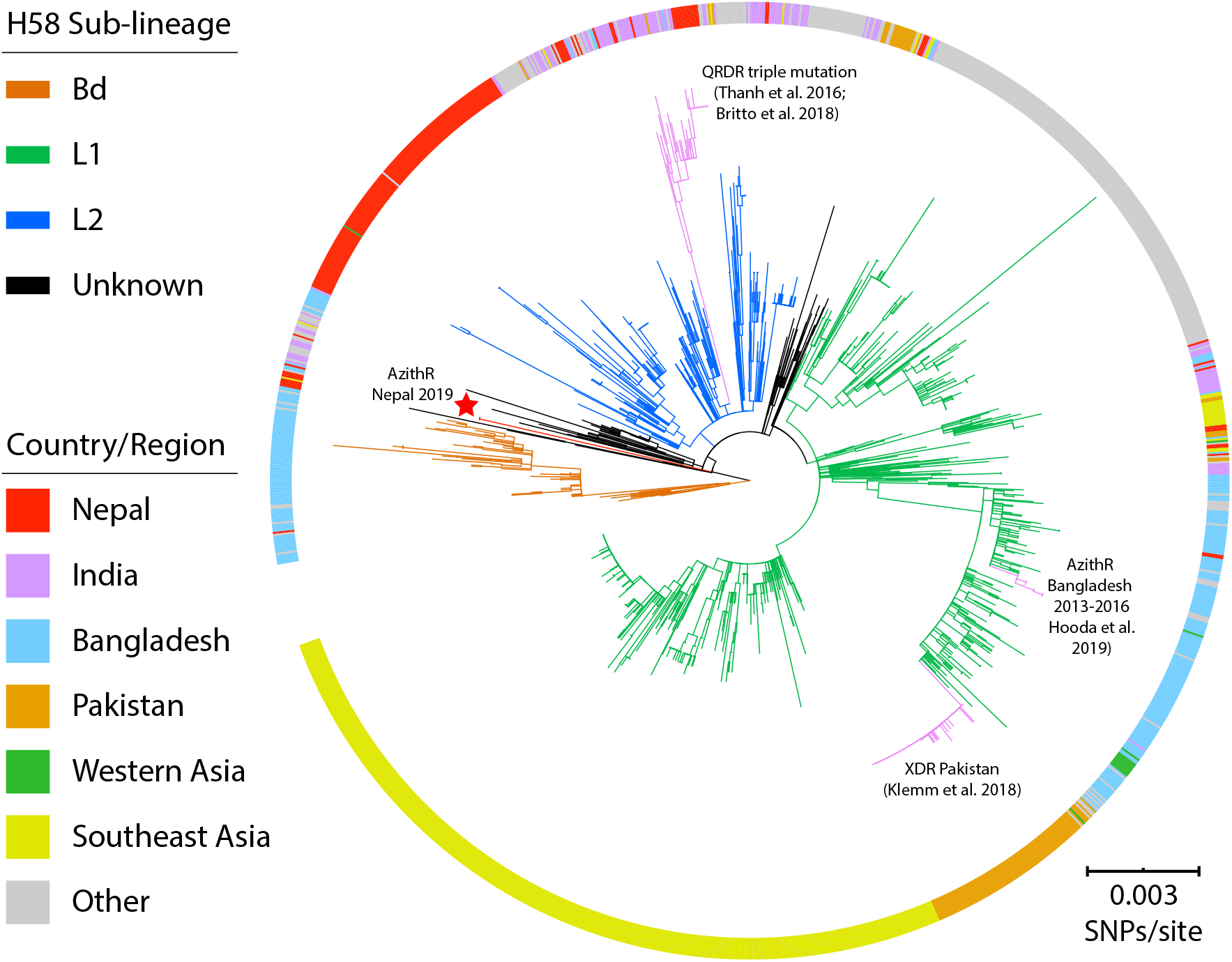
Phylogenetic relationships between azithromycin-resistant Nepali H58 isolates and global H58 isolates. Rooted maximum likelihood tree (CT18 was used as an outgroup to root the tree and pruned for visualization) reconstructed based on the SNPs of 1512 H58 isolates. Branches indicating three major H58 sub-lineages are colored in orange (sub-lineage Bd), green (sub-lineage I), and blue (sub-lineage II). The red terminal branch (also highlighted with a red star) shows the three azithromycin-resistant isolates described in this study. Branches highlighted in pink show different clusters associated with fluoroquinolone resistance, extensive drug resistance, and azithromycin resistance reported recently in Nepal, Pakistan and Bangladesh, respectively. The ring around the phylogeny indicates the location from which each isolate originates. The scale bar indicates 0.003 SNPS/site. QRDR: quinolone resistance-determining regions.

The emergence of Azith^R^ *S*. Typhi warrants further detailed clinical investigation to better understand the correlation between *in vitro* azithromycin susceptibility and clinical responses to azithromycin treatment. *In vitro* resistance to azithromycin does not often agree with *in vivo* effectiveness because the drug is mostly concentrated intracellularly.^17^ A previous study found no difference in response to azithromycin treatment among patients infected with S. Typhi exhibiting azithromycin MICs from 4-16 μg/ml.^18^ However, *S*. Typhi isolates with azithromycin MICs > 16 μg/ml are rare and clinical data on treatment responses in such organisms are lacking. Here, we report three patients infected with Azith^R^ *S*. Typhi with MIC >256 μg/ml, one of whom was given oral azithromycin 1g once/day. Though patient adherence to initial treatment cannot be ensured, clinical investigation suggests that the patient might not have adequately responded to azithromycin treatment and experienced microbiological failure. Further clinical and epidemiological investigations are needed to examine the increase of azithromycin resistance in S. Typhi in South Asia, limit its transmission, and improve empiric antimicrobial regimes.

Our data show that azithromycin resistance mutations at codon 717 *(acrB* gene) have independently emerged across distantly related lineages, suggesting that increasing use of azithromycin for treating typhoid fever may impose a strong selective pressure driving the emergence and spread of Azith^R^ *S*. Typhi. Convergent evolution towards azithromycin resistance in S. Typhi is of particular concern, as the development of azithromycin resistance mutations in XDR *S*. Typhi will eventually result in potentially untreatable infections. Further studies are needed to understand the driving forces and fitness effects of such resistance mutations. Typhoid conjugate vaccine (TCV) should be introduced in Nepal and other endemic countries to reduce the selective pressures induced by antimicrobial usage.

All three Azith^R^ *S*. Typhi isolates in this study were genetically identical, phylogenetically unrelated to all contemporary Nepali *S*. Typhi, and exhibited double mutations in the *gyrA* gene. We hypothesize these patients may have been infected from the same source. Additionally, the infecting organisms originate from an H58 variant that has been replaced by the dominant H58 sub-lineage II.^9^ This observation suggests this variant may be circulating at a low prevalence in the human population or have been maintained in a reservoir separate from the environmental transmission cycle, such as through asymptomatic carriage in the gallbladder. These data suggest these patients could have been infected from a chronic carrier within whom the infecting *S*. Typhi has evolved resistance to azithromycin.

In conclusion, we report three outpatients infected with highly azithromycin-resistant (MIC >256 μg/ml) S. Typhi in Nepal. One patient was given oral azithromycin 1g once/day and experienced prolonged fever until rescue treatment with ceftriaxone. All three Azith^R^ isolates were genetically identical H58 variants that were phylogenetically distinct from other contemporaneous Nepali *S*. Typhi. Azithromycin resistance was mediated by a chromosomal mutation R717L in the *arcB* gene. Further clinical and epidemiological investigations are required to track their transmission and evaluate clinical responses in patients infected with these important pathogens.

## Data Availability

All raw sequence data obtained from the study are deposited in ENA, project PRJEB37899.

## Acknowledgments

We would like to thank all members of the Molecular Epidemiology group at Oxford University Clinical Research Unit (OUCRU) in Vietnam; Microbiology Laboratory at Patan Hospital and doctors, nurses and health assistants of Patan Hospital Emergency and Outpatient Clinics.

## Author Contact Information

Pham Thanh Duy, Hospital for Tropical Diseases, Oxford University Clinical Research Unit, Ho Chi Minh City, Vietnam and Centre for Tropical Medicine and Global Health, University of Oxford, Oxford, United Kingdom. Sabina Dongol, Oxford University Clinical Research Unit, Patan Academy of Health Sciences, Kathmandu, Nepal. Abhishek Giri, Patan Hospital, Lalitpur, Kathmandu, Nepal. Nguyen Thi Nguyen To, Hospital for Tropical Diseases, Oxford University Clinical Research Unit, Ho Chi Minh City, Vietnam. Ho Ngoc Dan Thanh, Hospital for Tropical Diseases, Oxford University Clinical Research Unit, Ho Chi Minh City, Vietnam. Nguyen Pham Nhu Quynh, Hospital for Tropical Diseases, Oxford University Clinical Research Unit, Ho Chi Minh City, Vietnam. Pham Duc Trung, Hospital for Tropical Diseases, Oxford University Clinical Research Unit, Ho Chi Minh City, Vietnam. Guy Edward Thwaites, Hospital for Tropical Diseases, Oxford University Clinical Research Unit, Ho Chi Minh City, Vietnam and Centre for Tropical Medicine and Global Health, University of Oxford, Oxford, United Kingdom. Buddha Basnyat, Oxford University Clinical Research Unit, Patan Academy of Health Sciences, Kathmandu, Nepal. Stephen Baker, Cambridge Institute of Therapeutic Immunology & Infectious Disease (CITIID) Department of Medicine, University of Cambridge, Cambridge, UK. Maia Anita Rabaa, Hospital for Tropical Diseases, Oxford University Clinical Research Unit, Ho Chi Minh City, Vietnam and Centre for Tropical Medicine and Global Health, University of Oxford, Oxford, United Kingdom. Abhilasha Karkey, Oxford University Clinical Research Unit, Patan Academy of Health Sciences, Kathmandu, Nepal.

